# Predicting immune protection against outcomes of infectious disease from population-level effectiveness data with application to COVID-19

**DOI:** 10.1101/2024.10.17.24314397

**Authors:** Tianxiao Hao, Gerard E. Ryan, Michael J. Lydeamore, Deborah Cromer, James Wood, Jodie McVernon, James M. McCaw, Freya M. Shearer, Nick Golding

## Abstract

Quantifying the extent to which previous infections and vaccinations confer protection against future infection or disease outcomes is critical to managing the transmission and consequences of infectious diseases.

We present a general statistical model for predicting the strength of protection conferred by different immunising exposures (numbers, types, and variants of both vaccines and infections), against multiple outcomes of interest, whilst accounting for immune waning. We predict immune protection against key clinical outcomes: developing symptoms, hospitalisation, and death. We also predict transmission-related outcomes: acquisition of infection and onward transmission in breakthrough infections. These enable quantification of the impact of immunity on population-level transmission dynamics. Our model calibrates the level of immune protection, drawing on both population-level data, such as vaccine effectiveness estimates, and neutralising antibody levels as a correlate of protection. This enables the model to learn realised immunity levels beyond those which can be predicted by antibody kinetics or other correlates alone.

We demonstrate an application of the model for SARS-CoV-2, and predict the individual-level protective effectiveness conferred by natural infections with the Delta and the Omicron B.1.1.529 variants, and by the BioNTech-Pfizer (BNT162b2), Oxford-AstraZeneca (ChAdOx1), and 3rd-dose mRNA booster vaccines, against outcomes for both Delta and Omicron. We also demonstrate a use case of the model in late 2021 during the emergence of Omicron, showing how the model can be rapidly updated with emerging epidemiological data on multiple variants in the same population, to infer key immunogenicity and intrinsic transmissibility characteristics of the new variant, before these can be directly observed via vaccine effectiveness data.

This model provided timely inference on rapidly evolving epidemic situations of significant concern during the early stages of the COVID-19 pandemic. The general nature of the model enables it to be used to support management of a range of infectious diseases.

## 1 Introduction

Immune landscapes against infectious diseases are complex but vital to understand for infectious disease management [1]. The level of immune protection against an infectious disease conferred to an individual by vaccines and previous infections depends on many factors, including: the disease outcomes to be protected against (e.g., the likelihood of acquiring an infection or progression to severe disease), the source of immunity (differing by the type of vaccine or the variant of an natural immunising exposure), time since immunising exposure, and the variant of the infecting pathogen against which an immune response must be mounted. The ability to predict the level of protection from a combination of these factors for a real-world population helps inform public health response strategies, including designing vaccination programmes to achieve reduction targets in both mortality and morbidity burdens and in community transmission [2–5]. As most recently demonstrated by experiences with the COVID-19 pandemic, this ability to quantify the population-level impact of immunity from natural infections or from vaccination programmes is also critical for assessing if and when economically and socially costly [6–12] non-pharmaceutical interventions, such as lock-downs, can be relaxed without compromising public health goals [3, 13].

The level of protection from specific sources against specific outcomes at specific time-since-immunisation can be empirically observed through two sources: 1) vaccine *efficacy* trials, e.g., [14–18]; and 2) observed protection *effectiveness* from convalescent and vaccinated real-world populations, e.g., [19, 20]. Since efficacy and effectiveness data are rarely available for all combinations of outcome, source, and time since immunisation, a modelling framework is required to collate and standardise these sources of data, and to enable interpolation and extrapolation of protection, i.e., turning time-point estimates of immune protection into time-continuous curves against multiple outcomes, which can then further be used as inputs to risk assessments, or scenario projections.

Work by Khoury *et al*. [21] and Cromer et al. [22] have shown that a measurable and commonly reported correlate of protection (i.e., titres of neutralising antibodies against SARS-CoV-2) can be a useful intermediary quantity for 1) standardising efficacy estimates across clinical trials to enable comparison of efficacy across different sources of immunity, and 2) for generating continuous curves of predicted efficacy against various outcomes. However, only using clinically measured correlates of protection to predict efficacy has some limitations. First, modelling the level of immunity from one correlate of protection does not capture other mechanisms of immune response. For example, although titres of neutralising antibodies are identified as an effective correlate of protection against some outcomes from diseases such as influenza [23] and COVID-19 [21, 22, 24, 25], they only represent one aspect of the overall immune response, and other immune mechanisms, such as T-cells, make different contributions to immunity whose durability over time is not necessarily correlated with neutralising antibody levels [24–27]. This means that predicting immunity purely based on neutralising antibodies kinetics would likely underestimate long-term immunity provided by more enduring T-cells. Second, clinically measured vaccine efficacy is often an imperfect reflection of real-world effectiveness, limited by how representative (of real-world populations) the immune responses of the study cohort are, although collecting real-world effectiveness data has its own challenges too. Third, clinical efficacy and observational effectiveness data may be available for different combinations of immunising exposures and infecting pathogens/variants. Finally, rare but significant outcomes of public health importance, including mortality, are difficult to observe in clinical trials.

To improve upon an efficacy model parameterised on clinical measures of correlates of protection, we demonstrate in this work an expanded modelling framework incorporating both vaccine efficacy data and observational population-level effectiveness estimates. The ability to use both sources overcomes the aforementioned limitations of a clinical-efficacy-only model.

Here we present the use of the model for COVID-19 to demonstrate its methodology and utility in policy-making, and make available the code for adapting the model for other diseases, vaccines, and outcomes of interest. In our COVID-19 case study, we base our work on the model in Khoury *et al*.and Cromer *et al*.which links the degree of protection to titre counts of neutralising antibodies. However, rather than predicting directly from levels of neutralising antibodies, we predict the levels of protection from a latent quantity which we refer to as *immunity level*. We assume immunity level decays from its peak with a shape resembling antibody kinetics, but we calibrate the level of protection conferred by immunity levels with population-level effectiveness observations. Using the late-2021 detection and global spread of the Omicron variant as a case study on emerging variants, we also demonstrate how emerging data on variant-differentiated reinfection and effective reproduction rates can be incorporated as parameters in the model to infer possible ranges of key epidemiological parameters. Namely, we predict the level of immune evasion of the Omicron variant relative to the Delta variant and the Omicron basic reproduction number. This model extension allows us to rapidly estimate the likely levels of protection against outcomes of an Omicron infection, prior to the availability of any direct effectiveness and efficacy estimates.

## 2 Methods

### 2.1 Immune protection model definition

Our model assumes that the level of immune protection (immune effectiveness hereafter) against a pathogen conferred by vaccines and natural infections can be predicted from a latent parameter representing the level of immunity against the pathogen. We refer to these latent parameters, for different levels of existing immunity, as ‘immunity levels’. The decay of this latent parameter over time since peak immunity is set to follow the general shape of decay of clinically-measured correlates of protection from studies such as Khoury *et al*. [21], but the specific values of immunity level at different times since peak immunity are calibrated to match observational effectiveness data.

We assume that each immune individual *i* in a population has some immunity level *n*_*i,v*_ to variant *v*. An individual’s immunity level is assumed to be drawn from a normal distribution with mean *µ*_*s,d,v*_ based on the source of their immunity *s* (which may differ by vaccine dose or product, or in the case of natural immunity differ by infecting variant, or combinations thereof), the number of days *d* post peak immunity (i.e., the degree of waning), and the variant to be protected against *v*, and variance *σ*^2^, giving the inter-individual variation in immunity levels, which we assume to be constant across variants, sources of immunity and levels of waning (Equation 1).

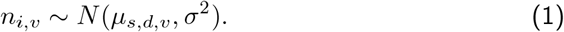

For each individual and for each type of outcome *o*, be that acquisition of infection, death, hospitalisation, symptomatic infection, or onward transmission, the probability that the outcome is averted, *E*_*o*_, is given by a sigmoid function, parameterised by a threshold immunity level *T*_*o*_, at which 50% of outcome events of type *o* are prevented, and a slope parameter *k* determining the steepness of this relationship (Equation 2). These parameters are assumed to be independent of the variant to be protected against and the source of immunity, which enables prediction of immune effectiveness to new situations such as new disease variants.

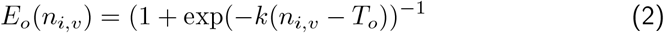

At the population-level, the immune effectiveness from a given source against a given outcome from a variant in a cohort with mean immunity level *µ*_*s,d,v*_ is the average probability over the whole population of the outcome being averted, *P*_*s,d,o,v*_. The population-level immune effectiveness as a function of immunity level is computed by integrating the sigmoid function with respect to the normal distribution of immunity levels (Equation 3).

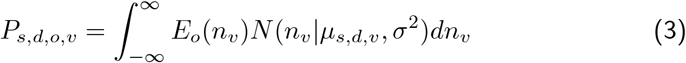

This integral has no known closed form and so a numerical approximation is computed in our implementation by Gauss-Legendre quadrature [28].

The mean immunity level, *µ*_*s,d,v*_ for a cohort with immunity source *s* and number of days *d* since peak immunity from that source is assumed to decay exponentially with time-since-peak-immunity, with half-life *H* days, from a peak mean immunity level against that variant of 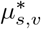 for each source (Equation 4)

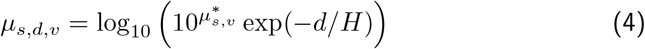

When extending the model to multiple variants, the peak mean immunity level against a given variant is then modelled as a log_10_ fold increase or decrease in immunity level of that variant, relative to an index variant (Equation 5).

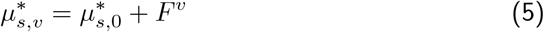

where for the index variant *F*^*v*^ = 0, for a variant with level of immune evasion relative to the index, *F*^*v*^ *<* 0, and for a variant more susceptible to immune protection than the index, *F*^*v*^ *>* 0.

Similar to the way we model variants, the peak immunity levels of different doses of the same vaccine are also modelled with log10 fold increase *G*^*booster*^ or decrease *G*^*single*^ in immunity level relative to the standard schedule (Equation 6).

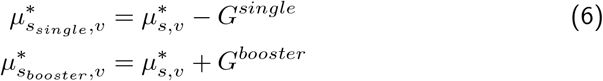

where 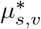 is the peak immunity level of the standard schedule, and *G*^*single*^ and *G*^*booster*^ are constrained to be non-negative, enforcing the level of immunity to be a monotonically increasing function of the number of doses. This allows us to use additional effectiveness estimates from studies that include partial first doses [29,30] and third ‘booster’ doses [19] in addition to standard schedules. We assume the increase in immunity level as doses increase is consistent across vaccine types, as there are typically insufficient data to parameterise them separately.

The decision to model peak immunity levels against different variants and from different vaccine dose numbers as relative to an index variant and a standard dose number enables the model to share information across different pairs of dose numbers and infecting variants. Importantly, this allows the model to e.g., predict protection against a new variant from estimates against an index variant and the estimated relative change parameter *F*^*v*^.

To fit the model to observational vaccine effectiveness estimates, we complete the model with a likelihood term. For vaccine effectiveness estimates provided as a point estimate and confidence interval, we define the the likelihood for vaccine effectiveness estimate *j* as a normal distribution over the logit-transformed estimate *V E*_*j*_, with mean given by the logit-transformed predicted effectiveness for that combination of source, days post-peak-immunity, outcome, and variant *P*_*s*_*j,dj,oj,vj* and with variance given by the sum of the square of the standard error of the estimate on the logit scale logit-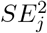 (approximated from provided uncertainty intervals of the source data), and an additional variance term 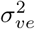, to represent any additional errors in these estimates arising from the observation process (Equation 7).

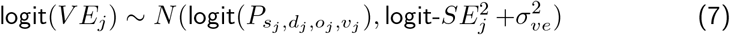

### 2.2 Model applications

#### 2.2.1 Fitting to vaccine effectiveness data against SARS-CoV-2

We demonstrate application of the model to population-level effectiveness estimates available in late 2021 of partial one-dose and primary two-dose course of the BioNTech-Pfizer (BNT162b2; Pfizer hereafter) and Oxford-AstraZeneca (ChAdOx1; AstraZeneca hereafter) vaccines against clinical outcomes of infections by the Delta variant of SARS-CoV-2 (death, severe disease, symptomatic infection) [20], and to estimates against acquisition of Delta infection (both symptomatic and asymptomatic), and onward transmission of breakthrough Delta infections [29, 30].

We then demonstrate an extended model, fitted to these data and to new effectiveness data available in early 2022 [19] of third-dose ’booster’ mRNA vaccines against symptomatic Delta infections, and to estimates of two-dose Pfizer and AstraZeneca, and 3rd dose booster vaccines against hospitalisation and symptomatic infections by the Omicron B.1.1.529 (referred to as Omicron hereafter) variant of SARS-CoV-2.

Note that because the only data for booster dose effectiveness available at the time of model fitting in late 2021 were of Pfizer boosters [19], we assumed that Pfizer booster immunity level is representative of any mRNA-based 3rd dose, irrespective of booster vaccine brand or the individual’s vaccine history. Finally, while immunity from natural infections is not represented in these vaccine effectiveness estimates, they can be predicted by extrapolating from the relative levels of immunity across different sources estimated in Khoury *et al*.. Hence, we also predict 1) the level of protection conferred by convalescence from natural Delta/wild-type (assumed to confer the same degree of immunity against Delta) infections against outcomes of Delta infections, and 2) protection conferred by Omicron infections, on its own or in combination with vaccines, against future Omicron infections — this is of interest because Omicron is assumed to possess evasion against all modelled sources of immunity, except previous exposures to Omicron itself.

All vaccine effectiveness estimates used to fit the model in the application we describe reported vaccine effectiveness for individuals pooled over a short period of time rather than a single day, so *d*_*j*_ was taken as the midpoint of that period. Although we model decay exponentially, the brevity of these time periods meant that using midpoints is an acceptable approximation.

#### 2.2.2 Rapid analysis of immune evasive variants: a case study on Omicron

When the Omicron variant first emerged as a public health concern in late 2021, early risk assessment and response planning were hindered by a lack of estimates on two key characteristics of Omicron: immunogenicity (i.e., the level of immune evasion possessed by Omicron, *F* ^O^, relative to the then-dominant Delta variant) and intrinsic transmissibility 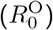. Importantly, there was also a lack of any datasets that could be used to estimate these key characteristics directly (e.g., neutralisation assays against Omicron to estimate *F* ^O^ or transmission studies in immune-naïve households to estimate 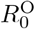), which only became available after widespread Omicron transmission globally.

To respond to these urgent inference needs in the earliest days of Omicron epidemic wave, we adapted the model to provide earliest available estimates on the immunogenicity and intrinsic transmissibility of Omicron, first using data that indirectly infers these quantities, then, as soon as they became available, using vaccine effectiveness and household transmission estimates on Omicron. The details of model adaptations, including additional parameters and uncertainties, data sources, and the chronological time of these methodological changes, are described in Appendix 1. Here, we provide a brief summary: we added *F* ^O^ and 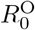 as latent parameters in the model. Then, taking advantage of the model’s ability to map between immune effectiveness for different variants, outcomes, sources of immunity, and degrees of waning, we fitted the model to the known intrinsic transmissibility of Delta, and a range of emerging data informing the relative immunogenicity and intrinsic transmissibility between the Omicron and the Delta variants. This allowed us to 1) jointly estimate *F* ^O^ and 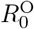, with appropriate representation of uncertainties in these data, and 2) predict immune protection against outcomes of Omicron infection.

### 2.3 Fitting the Model

Amongst model parameters,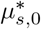, *H* and *σ*^2^ may either be inferred from clinical assays on correlates of protection, or calibrated against observational effectiveness data. The parameters *T*_*o*_ and *k* must be learned by fitting the model to effectiveness data. We keep the parameter *σ*^2^ fixed at the value estimated by Khoury *et al*.(0.465). We assign informative priors to 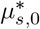, and *H* based on estimates from Khoury *et al*., and to *G*^*booster*^ based on estimates from UK Health Security Agency [19] after it had become available. The remaining parameters are given less informative priors. This enables the model to update these parameters based on the data, and fully incorporate uncertainty in these parameters into predictions. The prior distributions for all parameters in the main model are listed in Table 1 and additional parameters for the Omicron extension are listed in Appendix Table S1.

**Table 1:** Prior distributions for parameters as part of the immune effectiveness model. **Superscripts denote the range of values permitted for parameter distributions. ***Naïve implies that the priors are not informed by a specific literature source, but their distributions are selected to be relatively broad and fit to the data reasonably in prior simulations.

| Parameter | Prior distribution | Equation | Informed by | Description |
| --- | --- | --- | --- | --- |
| $k$ | $\mathcal{N}(0, 1)$ | 2 | Naïve*** | slope parameter in the logistic curve linking the probability of averting an infection outcome to an immunity level |
| $T_o$ | $\mathcal{N}(0, 1)$ | 2 | Naïve | threshold immunity level at which 50% of outcome events are prevented |
| $H$ | $\mathcal{N}(108, 10)$ | 4 | [21] | half-life of immunity level decay |
| $\mu_{AZ,\Delta}^*$ | $\mathcal{N}(-0.331, 0.0814)$ | 4 | [21] | standard two-dose schedule immunity levels (on $\log_{10}$ scale) of the AstraZeneca vaccine against the Delta variant at peak immunity, indexed on wild-type convalescent immunity level |
| $\mu_{Pfizer,\Delta}^*$ | $\mathcal{N}(0.327, 0.0775)$ | 4 | [21] | standard two-dose schedule immunity levels (on $\log_{10}$ scale) of the Pfizer vaccine against the Delta variant at peak immunity, indexed on wild-type convalescent immunity level |
| $G^{booster}$ | $\text{Lognormal}(5, 1)^{(0,\text{inf})**}$ | 6 | [19] | immunity level increase from a booster dose relative to a standard two-dose schedule |
| $G^{single}$ | $\mathcal{N}(1, 0)^{(0,\text{inf})}$ | 6 | Naïve | immunity level decrease relative to a standard two-dose schedule from a single dose of vaccine |
| $\sigma_{ve}^2$ | $\mathcal{N}(0, 1)^{(0,\text{inf})}$ | 7 | Naïve | additional variance term for observational vaccine effectiveness estimates |

All analyses were performed in R version 4.1.0 [31], using the greta package version 0.4.3 for model specification, inference, and predictions [32]. Inference was performed using 10 independent chains of Hamiltonian Monte Carlo, each run for 1000 samples after discarding 1000 warm-up iterations. Convergence was assessed by the potential scale reduction factor statistic (1.01 or less for all parameters), the effective sample size (greater than 500 for all parameters), and visual inspection of trace plots. Code used for this work is available at: https://github.com/idem-lab/neuts2efficacy.

The posterior distributions over immune effectiveness for different outcomes, vaccines, doses, and degrees of waning are computed to predict the range of immune effectiveness estimated for Delta and predicted for Omicron. Joint predictions of Omicron immune evasion and intrinsic transmissibility are also computed, for multiple iterations of the model fitted to different data, described in detail in Appendix 1.

## 3 Results

### 3.1 Immune protection predictions against the Delta variant

Figure 1 shows the estimated protection against the Delta variant over time since peak immunity, for various outcomes of interest: death, hospitalisation, symptomatic infection, acquisition of infection and onward transmission (given infection, i.e., breakthrough cases). The predicted immune effectiveness over time is consistent with the data (Figure 1), with the only notable difference being that the model predicts a slightly higher immune effectiveness from two-dose AstraZeneca vaccine against symptoms at around 100 to 150 days post peak compared to the immune effectiveness estimates from Andrews *et al*., 2021 [20].

**Figure 1:**
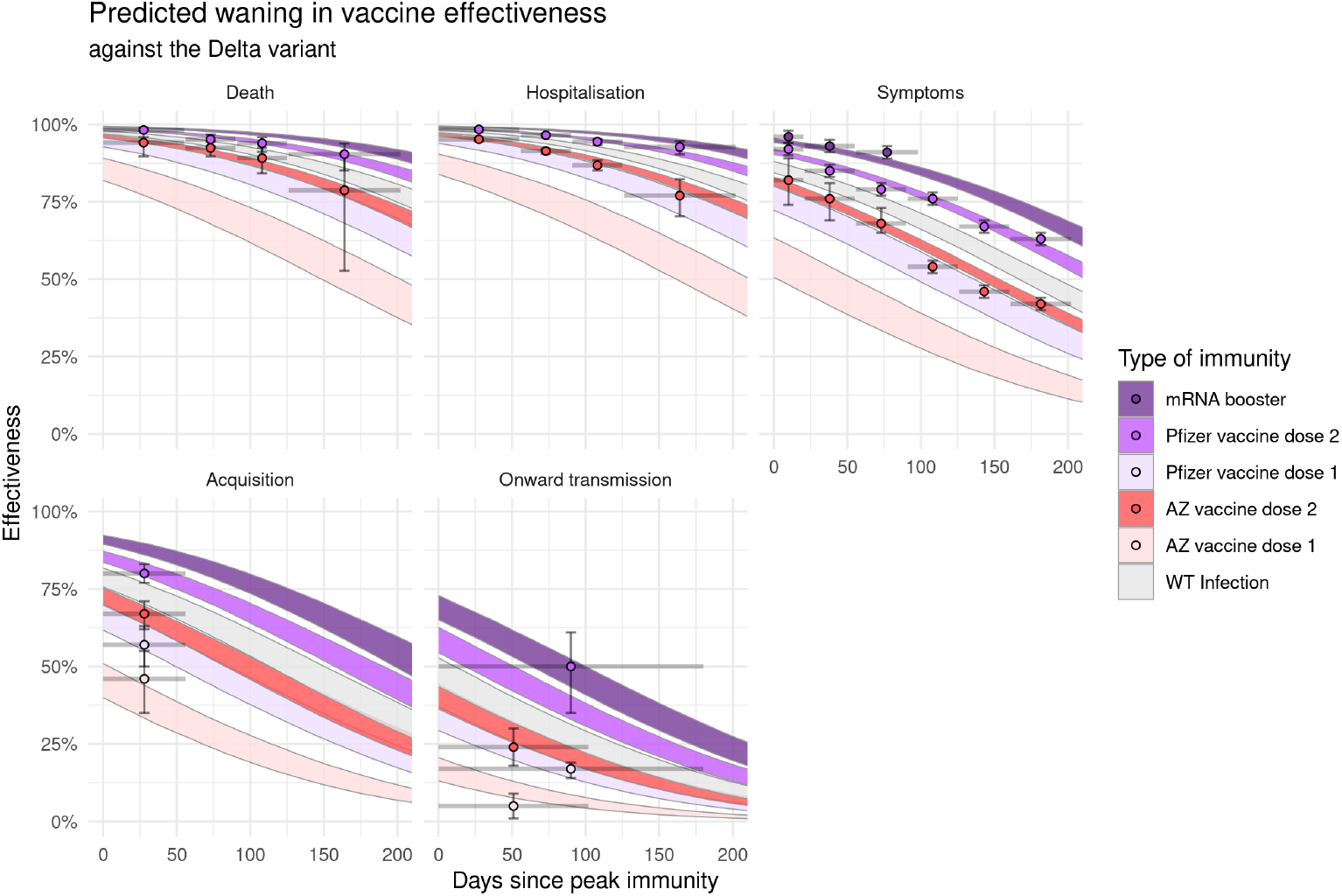
Estimated immune protection against the Delta variant over time since peak immunity (following administration of the first or second dose of either AstraZeneca (AZ) or Pfizer vaccines, or after boosting with an mRNA vaccine). WT = wild-type virus, which is assumed to confer the same level of immunity against the Delta variant as a Delta infection. Estimates of vaccine effectiveness from various observational studies are indicated by dots (point estimates) and vertical bars (95% confidence intervals), with horizontal bars indicating the range of days since peak immunity for individuals included in the study.

The model predicts that two doses of the mRNA Pfizer vaccine confer a higher protection than both two doses of the AstraZeneca vaccine and convalescence. Boosting with an mRNA product provides even higher protection than two doses of Pfizer vaccine, although the absolute differences in immune effectiveness is small for severe (death and hospitalisation) outcomes, since two doses of Pfizer vaccine already confers a very high degree of protection.

### 3.2 Predicting immune evasion and intrinsic transmissibility of Omicron from epidemiological data

The joint posterior distributions over the degree of immune evasion and the relative intrinsic transmissibility of the Omicron variant versus Delta (Figure 2) show how these estimated quantities changed as the model was re-parameterised with new, and increasingly informative, data. With only South African reinfection and reproduction rates data (left panel), the model estimated the Omicron variant to most likely be intrinsically less transmissible than the Delta variant, but possess a high degree of immune evasion (albeit with significant uncertainty on the strength of evasion). Incorporating the UK vaccine effectiveness estimates (middle panel) enabled the model to exclude very high levels of immune evasion from the plausible parameter space. Finally, when fitted to UK vaccine effectiveness estimates and Danish household attack rate data (right panel), the model estimated that Omicron is both intrinsically more transmissible, with a basic reproduction number around 1.2 times higher, and possesses a moderate to significant degree of immune evasion, capable of reducing immune effectiveness against transmission by 20-60% compared to the Delta variant. As the model was iteratively fitted to newer data, note that uncertainty in posterior distributions has decreased, although there was significant uncertainty even in the final iteration.

**Figure 2:**
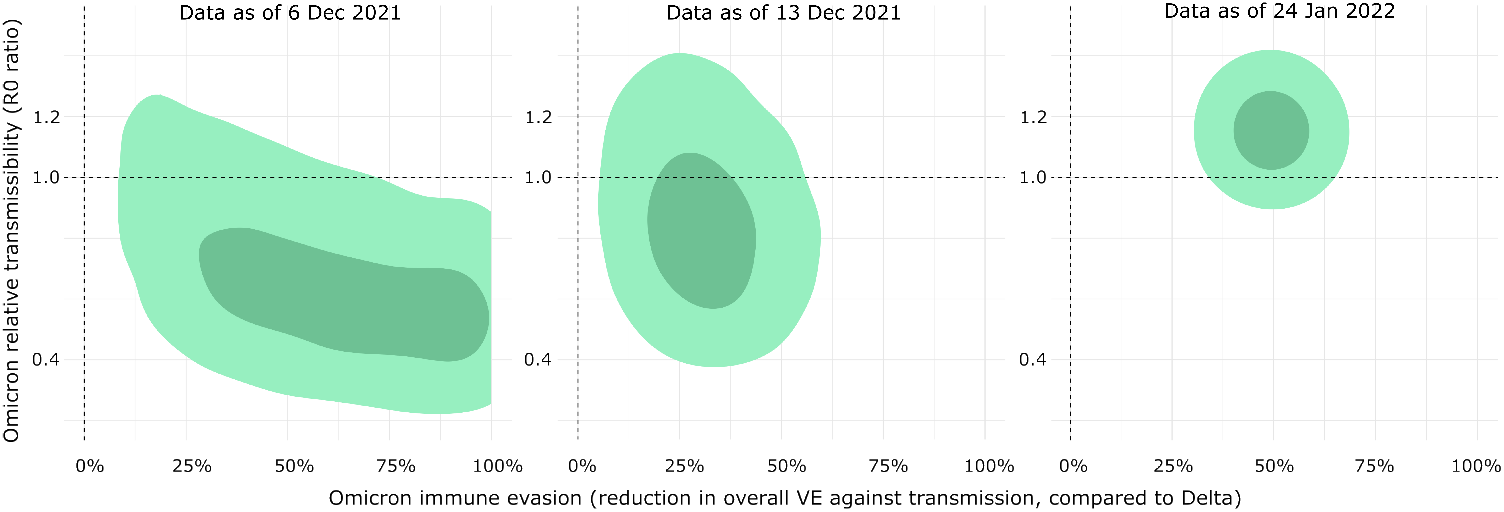
The joint posterior distributions over level of immune evasion and intrinsic transmissibility of the Omicron variant, relative to the Delta variant. The dark and light green areas show 95% and 50% density regions respectively. Joint parameter estimates are from three iterations of the model fitted to the latest data sources: left — South African reinfection and reproduction rates data, middle — South African data and UK vaccine effectiveness estimates, right — UK vaccine effectiveness estimates and Danish household attack rate data.

### 3.3 Immune protection predictions against the Omicron variant

Figure 3 shows predicted protection against the Omicron variant over time-since-peak-immunity, for various outcomes of interest, as predicted by the final iteration of the model in early 2022, including vaccine effectiveness data for both Delta and Omicron. Note that immune effectiveness predictions of single dose Pfizer and AZ vaccines are not shown here to assist visualising the more protective immunity profiles. When comparing the effectiveness predictions against symptomatic infections to the data in Andrews *et al*., 2022 [19] shown in Figure 3, the model is more pessimistic for two-dose AstraZeneca vaccines, especially in early phases of waning. The model is likewise more pessimistic for Pfizer effectiveness at peak protection, but it also predicts a more linear waning pattern compared to the data in Andrews *et al*., 2022, which appears more biphasic with more rapid decline in the first 100 days and minimal waning thereafter. For an mRNA booster, the model prediction against symptoms is consistent with the data up to around day 75, but it predicts slower waning compared to the data point around day 100. Note that the outlying data point for mRNA booster against symptoms at around day 150 was reported in the UK report at the time of analysis and thus was included in the example analysis representing this period of time, but it has since been removed in the published version of Andrews *et al*., 2022.

**Figure 3:**
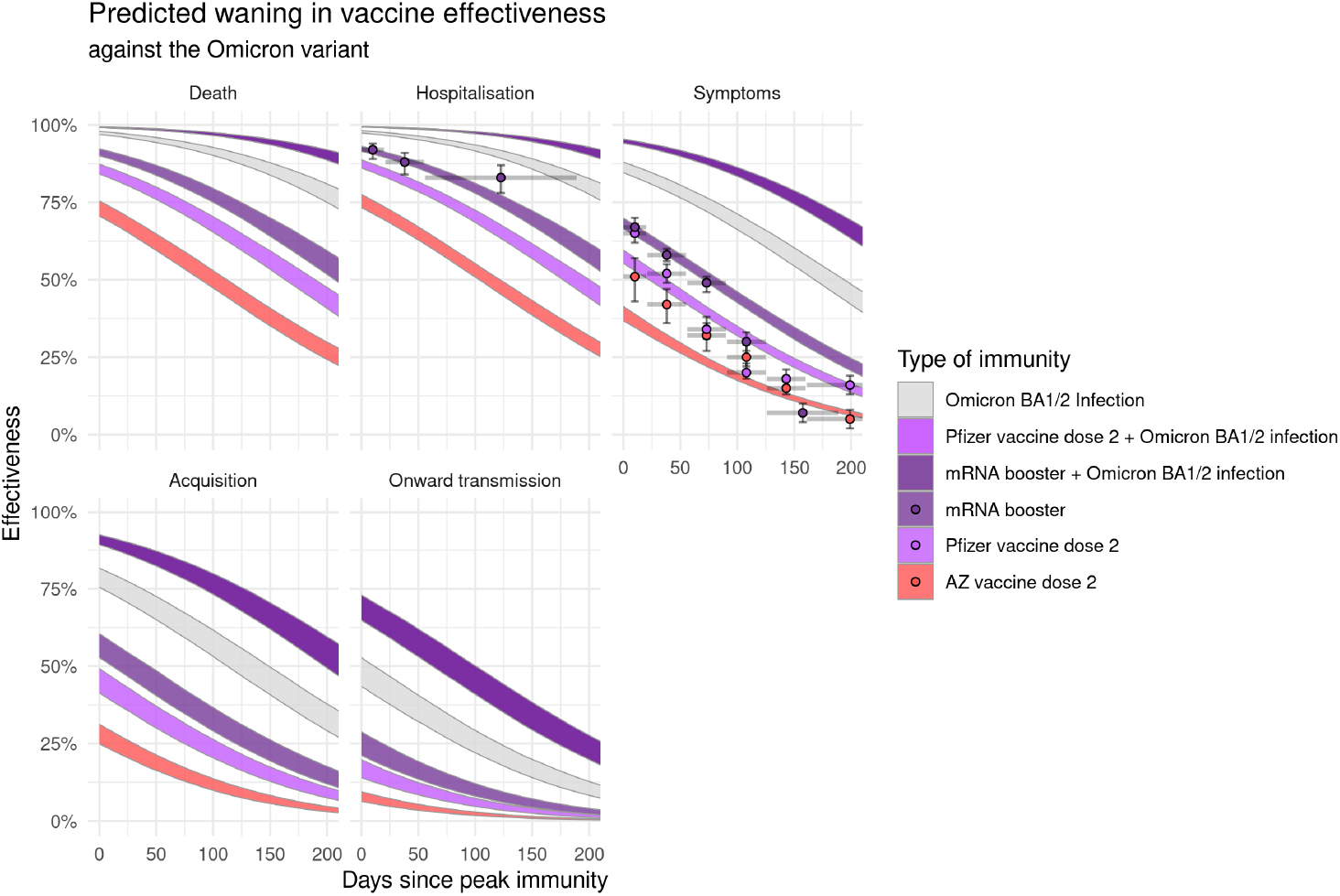
Estimated immune protection against the Omicron variant over time since peak immunity following administration of the second dose of either AstraZeneca or Pfizer vaccines, after boosting with an mRNA vaccine, and/or Omicron infection. Estimates of vaccine effectiveness from various observational studies are indicated by dots (point estimates) and vertical bars (95% confidence intervals), with horizontal bars indicating the range of days since immune event for individuals included in the study. Omicron sub-lineages BA.1 and BA.2 are assumed interchangeable both in the immune protection conferred by infections, and the level of protection against them.

Compared to immune effectiveness against the Delta variant, predicted immune effectiveness of mRNA booster, two-dose Pfizer, and two-dose AstraZeneca vaccines are all lower against the Omicron variant. However, the relative ranking of these three vaccine sources remains consistent, due to the assumption that the degree of immune evasion is constant across the modelled immunity sources. Infection with the Omicron variant confers greater immunity to subsequent Omicron exposure than any level of vaccination alone, due to the assumption that the level of immune protection from Omicron against Omicron reinfection is equivalent to that of previous wild-type infection against subsequent exposure to wild-type virus. Third dose (“booster dose”) combined with infection gives a further substantial increase in effectiveness, although at this point protection against severe outcomes cannot increase much further since it is close to 100% effective.

## 4 Discussion

The model presented in this work uses a combination of correlates of protection and vaccine effectiveness data to predict the level of protection conferred by vaccines and prior infections, against various outcomes of interest. Our model extends upon the neutralisation-level-to-clinical-efficacy model in Khoury *et al*. [21] by fitting the level of protection directly against observed real-world effectiveness data. Our approach is similar to that used by Hogan *et al*. [33], although the two models focus on different outcomes of infection. Hogan *et al*. focus on clinically relevant endpoints for evaluating vaccine effectiveness, including hospitalisation, death, and mild disease (mild symptomatic infection with some asymptomatic cases detected through routine screening). Among these, predicted protection against death is a particularly desirable addition to the outputs of Khoury *et al*. model as it has not been tested in clinical trials [22]. Our model also predicts protection against these outcomes, but by calibrating to a wider range of input data sources, our model additionally predicts the level of protection against all infections irrespective of clinical presentation and against onward transmission of breakthrough infections. This ability to predict protection against onward transmission is important because it can be combined with protection against acquisition of infection to calculate the reductive (time-dependent) effect of immunity on community transmission (as demonstrated in the Omicron extension in Appendix 1), which can be used in a range of subsequent analyses such as informing transmission parameters in dynamics models.

A powerful feature of our model is the flexibility to incorporate new types of parameters and data as needs arise. This was demonstrated in the Omicron adaptation case study detailed in Appendix 1 where we extend the model to fit to non-vaccine-effectiveness data (i.e., variant-differentiated reinfection and reproduction rates of Delta and Omicron and Omicron household secondary attack rates) and predict new outputs in addition to immune protection (i.e., immune evasion and intrinsic transmissibility of Omicron). By accurately representing the high degree of uncertainty in the new parameters, e.g., by assuming the proportion of the modelled South African population with immunity is between a broad range of 70-90% (the use of this parameter is described in detail in Appendix 1), we ensure the posterior estimations of the quantities of interest accurately reflect a large degree of uncertainty, given the indirect and limited nature fo these data sources for inferring immune responses. The flexibility of the modelling framework is also demonstrated in its ability to be rapidly updated in the Omicron case study, utilising newer and better data as they become available, to update risk assessments to inform public health policy. These evolving estimates were reported to the Australian Commonwealth government in near real-time, and helped inform key vaccine rollout policy changes during this critical period, which saw the largest COVID-19 wave in the country to date.

Our application of this model for epidemic analytics during the COVID-19 pandemic provided time-continuous and outcome-comprehensive predictions on the likely impacts of vaccine programmes. The model predictions showed that full-schedule vaccinations with either the Pfizer or AstraZeneca vaccines are effective against severe infection outcomes from both the Delta and Omicron variants of COVID-19, therefore demonstrating that vaccination is an effective tool for managing morbidity and mortality burdens. These findings are in broad agreement with similar estimates elsewhere [33, 34].

Before the widespread transmission of the immune evasive Omicron variant, a key aim of some COVID-19 vaccine programmes, in addition to alleviating mortality and morbidity burdens, was to achieve sustained reduction in community transmissions. This aim was for a time achieved in some populations such as in Australia in late 2021 [3, 35]. Important to this aim, our model predicts a fast rate of waning in protection against the two transmission-related outcomes of acquiring infections and onward transmissions. This suggests that the effectiveness of vaccine programmes in reducing transmission likely depends on repeated admissions of additional doses over time, broadly consistent with recommendations elsewhere [22, 36]. By linking protection against acquisition and onward transmission estimates from our model to information on the population-level immunity profile (proportions of the population with different types of immunity, from vaccination or prior infection), one can also predict reduction in community transmission due to immunity over continuous time or in hypothetical scenarios. An example of potential application is in Ryan *et al*. [3], where predicted reduction in transmission due to immunity is combined with vaccine uptake scenarios to simulate the levels of community transmission under different vaccination programme targets. An important finding of that study, which informed the Australian government’s vaccination uptake thresholds for national reopening of jurisdictional and international borders [35, 37], was determining the levels of vaccine coverage required to achieve targets in community transmission, and in turn whether and when non-pharmaceutical interventions could be relaxed, whilst still retaining control on transmission mainly through immunity. This potential use shows that the ability to estimate the reduction in transmission due to immunity is a major strength of our model with demonstrated wide-reaching policy impacts.

Due to its active use in risk assessment and policy planning objectives focusing on short-term impacts of immunity, our model was developed with an emphasis on short-term predictive accuracy. A further enhancement of the model would be to calibrate to effectiveness estimates over longer time since peak immunity, which were not available when the model was first developed. The model could also be reparameterised with a bi-phasic waning curve [33], where a more rapidly waning first phase corresponds to rapid changes in antibody kinetics post immunising exposure, and a more stable second phase reflecting when other more enduring mechanisms such as T-cells play a bigger role in immunity [27]. Implementing bi-phasic waning will likely bring the model’s prediction closer to the data for two-dose Pfizer vaccines against Omicron in Figure 3.

Another potential extension, should relevant data be available, is to model different levels of immune evasion in protection against different outcomes. For example, immune evasion could lead to increased transmissibility but not pathogenicity. The Omicron variant of SARS-CoV-2 demonstrates such a scenario, where mutations in spike epitopes of the virus enable it to evade neutralisation by antibodies and infect those with immunity, but the severity of these breakthrough infections is attenuated by other more robust mechanisms such as T cells [38, 39]. At the time of fitting our model, there was insufficient knowledge on the protection against severe outcomes from Omicron infections, so our model makes the necessary assumption of equal immune evasion against all outcomes, and learns effectiveness against Omicron mostly from the available data on symptomatic non-severe infections. Therefore, the model likely predicts a degree of immune evasion more consistent with non-severe outcomes (i.e., higher degree of immune evasion), likely explaining why the model under-predicted mRNA booster effectiveness against hospitalisation at around day 120 in Figure 3. With available future data, the interaction between immune evasion and outcomes can be explicitly modelled, for example with the addition of learned interaction coefficients *α*_*v,o*_ between pairs of variants and outcomes in Equation 5 (replacing the term *n*_*i,v*_ *− T*_*o*_ with *n*_*i,v*_ *− T*_*o*_ + *α*_*v,o*_).

As the code for implementing the model presented in this work is open source, it can be readily adapted for other research uses, such as modelling another disease, other vaccine types, or other outcomes of interest such as protection against developing ‘Long COVID’ (i.e., post-acute sequelae of COVID-19, [40]). A further avenue of research here is to develop the code-base into a software package with functions specifically designed to interface with a range of relevant data. This would increase the accessibility of the model to the pandemic preparedness research community, and enables its rapid deployment in public health response situations.

In conclusion, we present a model predicting the level of immune protection against various outcomes of infection using vaccine effectiveness data, and demonstrate its use for COVID-19. We showcase how the model outputs provide valuable and timely information on a range of epidemic response questions, and we further discuss potential improvements and extensions to the model that can be implemented with future data.

## Data Availability

All data used in this study are available at published sources:
https://www.medrxiv.org/content/10.1101/2021.09.28.21264260v2
https://www.nejm.org/doi/10.1056/NEJMoa2119451
https://www.medrxiv.org/content/10.1101/2021.09.15.21263583v2
https://www.nature.com/articles/s41591-021-01548-7
https://www.nature.com/articles/s41591-021-01377-8

## 5 Acknowledgements

This work received funding from the Department of Health and Aged Care Australia and the SPECTRUM Centre of Research Excellence funded by the Australian National Health and Medical Research Council (NHMRC; grant no. 1170960). T. Hao was supported by NHMRC Ideas Grant (grant no. 2019093), F. M. Shearer was supported by an NHMRC Investigator Grant (no. 2010051), D. Cromer was supported by an NHMRC Investigator Grant (no. 1173528).

