## Appendix 1 for "Predicting immune protection against outcomes of infectious disease from population-level effectiveness data with application to COVID-19"

### 1 Omicron model adaptation

In late 2021, we extended the model to provide rapid inference on the then-emerging Omicron variant of SARS-CoV-2. Particularly, we aimed to infer the range of plausible values for two key characteristics of Omicron: the immunogenicity (i.e., the level of immune evasion possessed by Omicron, $F^{\mathrm{O}}$, relative to the then-dominant Delta variant) and intrinsic transmissibility ($R_0^{\mathrm{O}}$).

Whilst the parameter $F^{\mathrm{O}}$ could be estimated directly from neutralisation assays against Omicron and $R_0^{\mathrm{O}}$ can be estimated from observed transmission events in immunologically-naïve populations, those were not available at the time of this case study in December 2021. We instead estimated these as a latent parameter and fitted the model to a range of data sources providing information on the two quantities of interest. Importantly, inferring $F^{\mathrm{O}}$ also enabled prediction of immune protection against clinical outcomes of Omicron infection. This suite of early estimates characterising plausible characteristics of Omicron were then available to inform simulation modelling of the likely consequences of an Omicron wave and impacts of potential interventions including bolstering vaccination programmes with additional doses.

#### 1.1 Fitting to epidemiological data from South Africa

When the earliest reports on epidemiological characteristics of the Omicron variant emerged in South Africa in the week commencing 6 December 2021, we expanded the model to define three likelihoods over three sources of epidemiological data:

1. estimates of the reinfection hazard ratio from Pulliam *et al.* [1] for both the Omicron emergence period and for the period of the Delta epidemic wave

2. estimates of the effective reproduction number of Delta in South Africa in late 2021 from a presentation by Carl Pearson, South African COVID-19 Modelling Consortium [2]

3. estimates of the ratio of the effective reproduction numbers of Omicron compared to Delta (as estimated from S-Gene Target Failure vs non-S-Gene Target Failure case counts, which was a widely used heuristic method for identifying Omicron cases at the time [3]) from the same presentation

Broadly, these three parameters each inform three different unknowns in the model: reinfection hazard ratios inform the degree of immune evasion of Omicron, relative to Delta; the reproduction number of Delta informs the degree of population immunity against this variant (since $R_0^{\Delta}$ estimates are available from other sources); and the ratio of reproduction numbers informs the intrinsic transmissibility of Omicron relative to Delta, after accounting for the degree of immune protection against each. The likelihoods for these three sources of data are detailed in turn below.

Note that all variables in the model that are uncertain and can influence the estimates of transmissibility or level of immune evasion are treated as unknown parameters, even if they are not quantities of interest or if they are not identified from the data. This enables integration over uncertainty in these parameters (i.e., a Monte Carlo simulation) whilst estimating parameters of interest.

#### 1.1.1 Reinfection hazard ratios

Pulliam *et al.* provide modelled estimates over time of the ratio between two hazards (rates of occurrence per unit time): the new infection hazard (rate of new infections per person without previous infection), and the reinfection hazard (rate of new infections among people who have had a previously reported infection). These estimates are informed by data on multiple infections from routine surveillance, and their model accounts for imperfect and differential case ascertainment between the two groups. They also provide an estimate of the ratio of these two hazards.

Assuming that the force of infection is the same for the two groups at the same point in time, the ratio of the reinfection hazard to the infection hazard can be interpreted as a proxy for the relative risk of infection between immune and non-immune individuals. This can further be interpreted as an estimate of one minus the effectiveness of prior immunity against *symptomatic* infection (symptomatic rather than any infection because the detection process is likely dependent on clinical presentation). Comparing the observed values of the reinfection-infection hazard ratio for late 2021 (approximately 0.3) and for the peak of the Delta epidemic wave in mid-2021 (approximately 0.1) against the values predicted by the immune effectiveness model for the dominant variants for a given $F^{\mathrm{O}}$ (and an assumed mean immunity level among those with immunity) enables quantitative inference about the degree of immune evasion of Omicron relative to Delta.

The likelihood for this data source is therefore given by Equation S1:

$$\begin{aligned}
\log(r_O) &\sim N(\log(\hat{r}_O), \sigma_r^2) \\
\log(r_\Delta) &\sim N(\log(\hat{r}_\Delta), \sigma_r^2) \\
\log(\hat{r}_O) &= \log(1 - P_{S,D,symptoms,O}) + \gamma \\
\log(\hat{r}_\Delta) &= \log(1 - P_{S,D,symptoms,\Delta}) + \gamma
\end{aligned} \tag{S1}$$

where $\log(r_v)$ denotes the log of the ratios given in [1], and $\log(\hat{r}_v)$ the log of the expected value of the reinfection hazard ratio during the period dominated by variant $v$ ($O$ indicating Omicron and $\Delta$ Delta), computed from the log of one minus the expected effectiveness of prior immunity against symptomatic disease, corrected by a log-offset parameter $\gamma$ to absorb any multiplicative bias in the hazard ratios (e.g., due to an incorrect specification of the ascertainment parameters in the model used to infer the hazard ratios). $P_{S,D,symptoms,v}$ is the modelled population-level effectiveness against symptomatic disease from variant $v$, given immunity source $S$, $D$ days since peak immunity. $P_{S,D,symptoms,\Delta}$ is defined by $\mu^*_{s,\Delta}$ — the baseline peak immunity level against Delta of an average immune person in the population (people with previous infection are the study population for these hazard ratios).

| Parameter | Prior distribution | Equation | Informed by | Description |
|---|---|---|---|---|
| $F^{\mathrm{O}}$ | $\mathcal{N}(0,1)$ | 5, S1 | Naïve | Omicron variant level of immune evasion parameter relative to Delta variant, though it does not directly appear in Equation S1, it underpins $P_{S,D,symptoms,O}$ |
| $\gamma$ | $\mathcal{N}(-1.26,0.18)$ | S1 | Naïve | correction factor of the reinfection hazard ratios, applied to both the Delta and Omicron periods in South Africa |
| $\mu^*_{s,\Delta}$ | $\mathcal{N}(0,1)^{(0,\mathrm{inf})}$ | S1, | Naïve | peak immunity immunity levels against the Delta variant from an average immune person in South Africa during late 2021. It underpins $P_{S,D,symptoms,\Delta}$ in Equation S1 |
| $D$ | $\mathcal{N}(120,20)$ | S1 | Naïve | days since peak immunity for an average immune person in South Africa during late 2021 |
| $Q$ | $\mathcal{N}(0.8,0.05)^{(0,1)}$ | S2 | Naïve | proportion of the South African population with immunity in late 2021 |
| $C$ | $\mathcal{N}(0.2,0.1)^{(0,0.5)}$ | S2 | Naïve | reduction in transmission achieved through behavioural changes for the South African population |
| $HSAR_\Delta$ | $\mathcal{N}(0.3,1)^{(0,1)}$ | S4 | [4] | household secondary attack rate of the Delta variant in a household with no immunity in Denmark in late 2021 |

Table S1: Prior distributions for additional parameters as part of Omicron extension to the immune effectiveness model.
**Superscripts denote the range of values permitted for parameter distributions.
***Naïve implies that the priors are not informed by a specific literature source, but their distributions are selected to be relatively broad and fit to the data reasonably in prior simulations.

$P_{S,D,symptoms,O}$ is additionally defined by the the Omicron level of immune evasion (relative to Delta) parameter $F^{\mathrm{O}}$. $\mu^*_{s,\Delta}$ is not calculated from a specific source of immunity, since the population-level immunity profile is difficult to reconstruct due to multiple past waves, imperfect ascertainment, and limited access to vaccination data. We instead treat $\mu^*_{s,\Delta}$ as unknown and specify it with a standard half normal prior to enforce that those with prior immunity must have had at minimum the level of immunity conferred by a single wild-type infection (Table S1). $D$ has a normal prior with mean of 120 days and standard deviation of 20 days, to represent the average time since infection in the Delta wave. The variance of the log reinfection-infection hazard ratio $\sigma_r^2$ is set to 0.25 to reflect a significant degree of uncertainty in the relationship between these heavily modelled estimates and the adjusted relative risks. This high uncertainty also allows immune protection against symptomatic infection (to which the model is calibrated) to be different from immunity against symptomatic reinfection (which we have no data to calibrate against). $r_O = 0.3$ and $r_\Delta = 0.1$, which were read off Figure 5 in Pulliam *et al.*.

#### 1.1.2 Delta Reproduction Number

Whilst the Delta variant was estimated to have the highest intrinsic transmissibility among identified variants prior to Omicron (estimates range from 6-8; [5]), estimates of the effective reproduction number in Gauteng Province, South Africa at the time of the Omicron outbreak were largely just below 1. There were minimal non-pharmaceutical interventions in place in Gauteng province during this time, and estimates of mobility from Google community mobility reports [6] indicate that population mixing rates were largely at or above baseline levels (with the exception of visits to workplaces at a 7% reduction). The majority of the reduction in transmission from $R_0$ conditions in Gauteng was therefore likely to have been due to prior immunity from previous epidemic waves, with some small additional effect of vaccination (vaccine coverage was low at the time). Given the multiple waves of infections with different variants (wild-type, Beta, Delta), and evidence of multiple re-infections over 2021 (see Figures 2–3 in Pulliam *et al.*), the average level of immunity in the population was likely substantially higher than that conferred by just a single Delta infection during the mid-2021 Delta wave, meaning that it could not be modelled assuming just one immunising exposure from a Delta infection. Instead, the degree of immunity was to be inferred from an assumed intrinsic reproduction number ($R_0^\Delta$) and estimate of the effective reproduction number ($R_{\text{eff}}^\Delta$) of Delta, given assumptions about the effective fraction of the population with any immunity ($Q$) and the effect of human behaviour, public health and social measures, and isolation of cases on transmission ($C$).

The likelihood on estimates of the reproduction number of Delta in Gauteng Province in November 2021 was therefore defined as in Equation S2.

$$\begin{aligned}
\log(R_{\text{eff}}^\Delta) &\sim N(\log(\hat{R}_{\text{eff}}^\Delta), \sigma_R^2) \\
\hat{R}_{\text{eff}}^\Delta &= R_0^\Delta \cdot (1 - C) \cdot I^\Delta \\
I_\Delta &= A_\Delta \cdot T_\Delta \\
A_\Delta &= Q\,(1 - P_{S,D,\text{acquisition},\Delta}) + (1 - Q) \\
T_\Delta &= Q\,(1 - P_{S,D,\text{transmission},\Delta}) + (1 - Q)
\end{aligned} \tag{S2}$$

$R_{\text{eff}}^\Delta$ is the effective reproduction number of Delta, the logarithm of which is normally distributed with mean $log(\hat{R}_{\text{eff}}^\Delta)$ and variance $\sigma_R^2$. We specified $R_{\text{eff}}^\Delta = 0.8$ and $\sigma_R^2 = 0.2$ following estimates by Pearson *et al.* [2]. $\hat{R}_{\text{eff}}^\Delta$ is calculated as the basic reproduction number multiplied by reduction due to behavioural effects $C$ and reduction due to immunity $I_\Delta$. $I_\Delta$ is calculated as the product of the population-level reductions due to immunity against acquisition of infection ($A_\Delta$) and against onward transmission of breakthrough infections ($T_\Delta$), each of which is calculated from the corresponding immune effectiveness against Delta, given the modelled level of peak immunity and time since peak (as in the previous section) and the fraction of the population with immunity $Q$.

$Q$ is assigned an informative normal prior with mean 0.8 and standard deviation 0.05 (70-90% have some immunity), truncated to the unit interval. We assumed that $R_0^\Delta = 6$ and that $C$ has a normal prior distribution with mean 0.2 (20% reduction in transmission from behaviour), a standard deviation 0.1, and truncated between 0 and 0.5 (maximum 50% reduction in transmission).

#### 1.1.3 Reproduction number ratio

Pearson *et al.*provides an estimate of the ratio of the effective reproduction number of Omicron to Delta in Gauteng Province, with quantification of uncertainty. The estimate is computed from timeseries of case counts with (assumed Omicron) and without (assumed Delta) S-gene target failure. The expected effective reproduction number for each variant can be computed as for $\hat{R}_{\text{eff}}^\Delta$ in Equation S2 and the ratio can be computed and a likelihood defined as follows:

$$\frac{R_{\text{eff}}^{O}}{R_{\text{eff}}^{\Delta}} \sim N\left(\frac{\hat{R}_{\text{eff}}^{O}}{\hat{R}_{\text{eff}}^{\Delta}}, \sigma_{R_{\text{eff}}}^{2}\right)$$
$$\frac{\hat{R}_{\text{eff}}^{O}}{\hat{R}_{\text{eff}}^{\Delta}} = \frac{R_0^{\text{O}}}{R_0^{\Delta}} \cdot \frac{I_O}{I_\Delta} \tag{S3}$$

where $R_{\text{eff}}^{\text{O}}/R_{\text{eff}}^{\Delta} = 2.1$ and $\sigma_{R_{\text{eff}}}^{2} = 0.41$ to match the distribution of estimates from Pearson (across estimates with the same and different generation intervals between variants). Note that reduction in transmission due to behaviour $C$ is assumed to be constant across variants thus it cancels out in this ratio.

### 1.2 Fitting to real-world vaccine effectiveness data against the Omicron variant of SARS-CoV-2

While the Omicron analysis described above was performed before any effectiveness estimates against Omicron were available, in the week commencing 13 December 2021, we updated the model to include the first batch of population-wide estimates on immune protection against Omicron, rapidly made available by the UK Health Security Agency — these estimates are now published in Andrews *et al.* [7]. These estimates provide observation on immune effectiveness against both Omicron and Delta for populations with the same immunity profile, thus allowing inference on the relative immune evasion of Omicron. Note the estimates for effectiveness of AstraZeneca vaccination against Omicron are highly uncertain and have negative point estimates, and the authors note that these estimates are likely to be untrustworthy due to the fact that AstraZeneca vaccinees are more likely to be vulnerable (and more prone to clinical disease). For this reason, estimates of effectiveness of the AstraZeneca vaccine were excluded, as were any effectiveness estimates computed on fewer than 10 Omicron cases (e.g. Pfizer dose 2 in the period immediately following vaccination).

These additional immune effectiveness estimates were used to inform the model with a likelihood defined as described in Equation 7 in the main text.

### 1.3 Incorporating household secondary attack rates

In addition to the aforementioned data sources,household transmission studies can provide key information about the characteristics of circulating variants.

In January 2022, Lyngse *et al.* [4] provided additional estimates on household secondary attack rates (HSAR) of Omicron compared to Delta in Denmark, we incorporated these estimates into the model in the week commencing 23 January 2022. Crucially, Lyngse *et al.*provided estimates on the odds ratio between Omicron and Delta HSAR in immune-naïve households (unvaccinated with no known infection history), which can be interpreted as the ratio between intrinsic transmissibility of the two variants, since there is no effect of immunity. These data provide more direct and certain inference on $R_0$ ratio compared to approach based on effective reproduction number ratios and a modelled effect of immunity in South African data, so we removed the South African data in Section S1.1, and re-parameterised the model based on UK vaccine effectiveness data (for inferring immune evasion) and Lyngse *et al.*HSAR data (for inferring intrinsic transmissibility).

Note that the definition of HSAR in Lyngse *et al.*more closely aligns with household final attack risk (FAR), due to the simplifying assumption that all in-household transmissions originate from one infector — see Sharker *et al.* [8] for a detailed explanation. We therefore convert these FAR reported in Lyngse *et al.*to true HSAR using Figure 1 in Sharker *et al.*, assuming linearity in the HSAR-FAR ratio over the range of FAR values. The observed $\frac{FAR_O}{FAR_\Delta}$ ratio in Lyngse *et al.*is defined with a likelihood in Equation SS4:

$$\begin{aligned} \frac{FAR_O}{FAR_\Delta} &\sim N(\frac{F\hat{A}R_O}{F\hat{A}R_\Delta}, \sigma^2_{FAR}) \\ F\hat{A}R_\Delta &= f(HSAR_\Delta) \\ F\hat{A}R_O &= f(HSAR_O) \\ HSAR_O &= HSAR_\Delta \cdot \frac{R_0^{\mathrm{O}}}{R_0^{\Delta}} \end{aligned} \tag{S4}$$

where $\frac{FAR_O}{FAR_\Delta}$ follows a normal distribution with mean $\frac{F\hat{A}R_O}{F\hat{A}R_\Delta}$ and variance $\sigma^2_{FAR}$. $F\hat{A}R_O$ and $F\hat{A}R_\Delta$ are estimated FAR for the two variants, and $f(\cdot)$ is a linear regression function with fixed parameters mapping between FAR and HSAR, based on Figure 1 in Sharker *et al.*. $HSAR_O$ is linked to $HSAR_\Delta$ via the $R_0$ ratio, and $HSAR_\Delta$ follows a prior normal distribution with mean of 0.3 and variance of 1. $\frac{FAR_O}{FAR_\Delta} = 1.17$ and $\sigma^2_{FAR} = 0.099$ as reported in Lyngse *et al.*(note the values are slightly different in the now published version).

### 1.4 Model outputs

Model fitting procedures and immune protection predictions are described in the main text. However, in addition, the posterior samples from each version of the model fit are used to visualise the joint posterior distribution over level of immune evasion and intrinsic transmissibility of Omicron, relative to Delta, through a kernel density estimate over pairs of parameter samples. The joint posterior distributions are visualised for three iterations of the model: South Africa data only, South Africa and UK immune effectiveness data, and UK immune effectiveness and Danish household data. Note that South African data was not included in the final model iteration as the combination of vaccine effectiveness and household transmission data meant that the population-level South African data contributed little to the posterior by this stage. The level of immune evasion is calculated as:

$$1 - \frac{(1 - I_O)}{(1 - I_\Delta)} \tag{S5}$$

but with $Q$ fixed at 1 when calculating $I_O$ and $I_\Delta$. That is, it is the reduction in the effect of immunity on transmission shown by Omicron, relative to Delta, for a population where all members have the same level of immunity.

[6] COVID-19 Community Mobility Report.

[7] Nick Andrews, Julia Stowe, Freja Kirsebom, Samuel Toffa, Tim Rickeard, Eileen Gallagher, Charlotte Gower, Meaghan Kall, Natalie Groves, Anne-Marie O'Connell, David Simons, Paula B. Blomquist, Asad Zaidi, Sophie Nash, Nurin Iwani Binti Abdul Aziz, Simon Thelwall, Gavin Dabrera, Richard Myers, Gayatri Amirthalingam, Saheer Gharbia, Jeffrey C. Barrett, Richard Elson, Shamez N. Ladhani, Neil Ferguson, Maria Zambon, Colin N.J. Campbell, Kevin Brown, Susan Hopkins, Meera Chand, Mary Ramsay, and Jamie Lopez Bernal. Covid-19 Vaccine Effectiveness against the Omicron (B.1.1.529) Variant. *New England Journal of Medicine*, 386(16):1532–1546, April 2022. Publisher: Massachusetts Medical Society.

[8] Yushuf Sharker and Eben Kenah. Estimating and interpreting secondary attack risk: Binomial considered biased. *PLOS Computational Biology*, 17(1):e1008601, January 2021. Publisher: Public Library of Science.
